# Dual-energy X-ray absorptiometry derived knee shape may provide a useful imaging biomarker for predicting total knee replacement: findings from a study of 37,843 people in UK Biobank

**DOI:** 10.1101/2024.01.04.24300833

**Authors:** Rhona A Beynon, Fiona R Saunders, Raja Ebsim, Monika Frysz, Benjamin G Faber, Jennifer S Gregory, Claudia Lindner, Aliya Sarmanova, Richard M Aspden, Nicholas C Harvey, Timothy Cootes, Jonathan H Tobias

## Abstract

**Objective:** We developed a novel imaging biomarker derived from knee dual-energy x-ray absorptiometry (DXA) to predict subsequent total knee replacement (TKR). The biomarker is based on knee shape, determined through statistical shape modelling. It was developed and evaluated using data and scans from the UK Biobank cohort.

**Methods:** Using a 129-point statistical shape model (SSM), knee shape (B-score) and minimum joint space width (mJSW) of the medial joint compartment (binarized as above or below the first quartile) were derived. Osteophytes were manually graded in a subset of DXA images. Cox proportional hazards models were used to examine the associations of B-score, mJSW and osteophyte score with the risk of TKR, adjusted for age, sex, height and weight.

**Results:** The analysis included 37,843 individuals (mean 63.7 years). In adjusted models, B-score and mJSW were associated with TKR: a standard deviation increase in B-score was associated with a hazard ratio (HR) of 2.32 (2.13, 2.54), and a lower mJSW with a HR of 2.21 (1.76, 2.76). In the 6,719 images scored for osteophytes, mJSW was replaced by osteophyte score in the most strongly predictive model for TKR. In subsequent ROC analyses, a model combining B-score, osteophyte score, and demographic variables had superior discrimination (AUC=0.87) in predicting TKR at five years compared with a model with demographic variables alone (AUC=0.73).

**Conclusions:** An imaging biomarker derived from knee DXA scans reflecting knee shape and osteophytes, in conjunction with demographic factors, could help identify those at high risk of TKR, in whom preventative strategies should be targeted.

## Introduction

Knee osteoarthritis (kOA) exhibits distinct radiological features, including osteophyte formation and joint space narrowing (JSN), the latter primarily affecting the medial joint compartment in primary kOA. Additionally, the condition is associated with characteristic shape alterations, including varus and valgus alignment, which can both result from and contribute to the disease (1–3).

Currently, Joint Space Width (JSW) is the only accepted biomarker for kOA progression in therapeutic trials (4, 5). However, grading systems such as the Kellgren-Lawrence system (6), which incorporates both osteophytes and JSN into a single severity scale, are often used in epidemiological studies. While the Kellgren-Lawrence grading system provides a valuable tool for defining kOA in research, it has limitations in predicting clinical outcomes, as evidenced by its weak association with pain and function (7, 8) and it’s limited sensitivity to change (9, 10). As a consequence, it is not commonly employed in clinical practice. Indeed, studies have shown that preoperative radiographs evaluated using the Kellgren-Lawrence system often underestimate the severity of kOA (11). This underscores the need to consider additional factors beyond the scope of the grading system, with knee shape emerging as a potential influential factor in the progression of the disease.

Statistical shape modelling (SSM), facilitated by machine learning techniques, holds the potential to enhance the prediction of osteoarthritis progression by identifying distinct joint shape features that are associated with adverse clinical outcomes. However, the application of this method in the context of the knee remains relatively unexplored, with existing studies primarily focusing on three-dimensional (3D) imaging modalities (12–14). For example, Bowes et al. developed the B-score to capture femoral shape changes in kOA on MRI images (14). Elevated B-scores were associated with a 60% increased risk of TKR and its predictive ability was comparable to that of the Kellgren-Lawrence grade.

DXA imaging is gaining interest for joint shape evaluation due to its advantages of low radiation exposure and cost, and widespread availability. Moreover, modern high-resolution DXA scanners generate high-quality images similar to radiographs, making it a viable option for screening individuals at high risk of osteoarthritis progression. Observed shape variations on hip DXA, including reduced acetabular coverage and cam morphology, have already been linked to advanced disease (15, 16) and statistical shape modes have identified changes in hip shape that are linked to disease progression (17). While the use of knee DXA scans in assessing kOA has not been extensively investigated, a study successfully used SSM to capture alterations in knee morphology in a small cohort of kOA patients through sequential DXA images spanning 6–12 months (10).

In this study, we aimed to investigate the potential utility of knee DXA scans as a screening tool for identifying individuals with kOA who are at a high risk of progressing to total knee replacement (TKR), based on evaluating knee shape. To achieve this, we developed and subsequently applied a statistical shape model to approximately 40,000 knee DXA scans from UK Biobank (UKB). We examined the relationship between knee shape, quantified by B-score, and risk of subsequent TKR. Additionally, we explored whether any predictive value of B-score could be improved by incorporating additional features obtained from knee DXA scans. Specifically, we examined the inclusion of minimum joint space width (mJSW) and osteophyte classification (available in a subset of approximately 7,000 scans) along with conventional risk factors, namely age, sex, height, and weight.

## Materials and methods

### Participants

We used data from the UKB extended imaging study (18), a large-scale research study that aims to collect and analyse medical imaging data, including DXA scans, from approximately 100,000 participants in the UKB cohort (19). At both baseline and imaging visits, UKB participants completed a touch-screen questionnaire, underwent a nurse-lead interview, and had physical measurements taken. Further information was obtained via data linkage, including hospital episode statistics (HES) (20).

All subjects provided written informed consent before participation. UKB has full ethical approval from the National Information Governance Board for Health and Social Care and the North-West Multi-Centre Research Ethics Committee (11/NW/0382). Permission to access and analyse UKB data for this study was approved under UKB application number 17295.

### Acquisition of Knee DXA images

DXA scans were acquired using a Lunar iDXA scanner (GE Healthcare), with participants in a non-weight bearing supine position (21). High-resolution DICOM format images were downloaded from the UKB showcase (downloaded in April 2021). Individuals with prior TKR were excluded from the analysis.

### Ascertainment of outcomes

Our primary outcome of interest was TKR. Additionally, we examined cases of hospital-diagnosed knee osteoarthritis (HES-kOA), as a secondary outcome. Both outcomes were identified via linkage to the HES database (see Supplementary Table 1 for a list of the diagnostic codes). All UKB participants were linked, both prospectively and retrospectively at baseline. Records begin on April 1st, 1997, and the data were downloaded in October 2022 (capturing information up until the end of December 2021).

### Assessment of covariates

Participants’ height and weight were measured prior to imaging using standardised procedures (22). Age and sex were collected at the time of enrolment into UKB and were self-reported.

### Statistical shape modelling

The SSM techniques employed in this study have been described elsewhere (23, 24) and additional information is provided in the Supplementary Methods. SSM was performed using software developed at The University of Manchester (25). A 129-point template was developed, covering the left distal femur, proximal tibia, proximal fibula, and superior patella, and excluding any osteophytes (Supplementary Figure 1). An automatic BoneFinder® (26) search model was trained (using 6,718 images from an initial pool of 7,000 images) to apply the template to new images. The accuracy of the search model was evaluated using 3-fold cross-validation experiments (Supplementary Table 2). The BoneFinder® model was applied to the remaining left knee DXA images (n=31,207; see Supplementary Methods) to automatically place the points. When necessary (for 4,214 images), trained annotators (RB and FS) manually corrected point placements to improve the precision of the SSM (Supplementary Table 3).

The first 27 statistical knee shape modes (KSMs) obtained from SSM were standardized using the sample standard deviation. To limit multiple comparisons, we focused on the first 10 KSMs, explaining 79.5% of shape variance. Subsequent KSMs each made minimal contributions to the sample variance (≤2%) (Supplementary Figure 2).

### Generation of a quantitative measure of knee shape: B-score

A knee shape variable was created using all KSMs from the SSM. A vector representing the line connecting the mean knee joint shape of a “healthy” population and that of a “diseased” population was generated for each outcome. The KSMs for each image were projected onto this vector to obtain the B-Score (14). Each B-score unit represents one standard deviation (SD) from the mean knee joint shape of the healthy population (assigned a B-score of 0). As well as calculating B-scores for the entire population, sex-specific B-scores were derived for sensitivity analysis (Supplementary Methods).

### DXA-based measures of joint space and osteophytes

A custom script automatically measured mJSW of the medial and lateral compartments using specific template points on the distal femur and proximal tibia (Supplementary Methods). Medial mJSW values were divided into quartiles, and a binary variable was generated to compare the first quartile against the second, third and fourth.

Osteophytes were assessed in a subset of 6,719 DXA images, selected from the initial pool of 7,000 images designated for the search training set (see Supplementary Methods). Using a DXA-based reference Atlas (Supplementary Document), we initially conducted visual grading on a 0-3 scale. We then computed cumulative values (ranging from 0 to 12) by aggregating grades from all four sites on the medial and lateral aspects of the femur and tibia. An osteophyte score was then assigned based on the total sum: 0 (sum = 0), 1 (sum = 1), 2 (sum = 2-3), 3 (sum = 4 or greater), as outlined in Supplementary Table 4.

### Statistical analysis

We employed Cox proportional hazards modelling and logistic regression to assess the relationships between KSMs, B-scores, mJSW, and osteophytes with the incidence of TKR and HES-kOA, respectively. The proportional hazards assumption was verified using the Schoenfeld residuals approach. Results are reported as hazard ratios (HRs) and odds ratios (ORs) with their corresponding 95% confidence intervals (CIs). Both unadjusted and adjusted analyses were conducted, with adjustments made for age, sex, height, and weight. Age, height, and weight were treated as continuous variables, while sex was considered as a binary variable. Where death occurred before TKR, the event was censored at the time of death.

To evaluate the predictive performance of the models for TKR and HES-KOA, we used the Harrell’s Concordance index (C-index) and area under the receiver operating characteristic curve (AUC), in combination with Akaike information criterion (AIC) and Bayesian information criterion (BIC). Initially, we ran univariate models for B-score, mJSW, and osteophyte score using the subset of participants with osteophyte data (n=6,719). We then examined a model incorporating the demographic factors age, sex, height, and weight as additional variables. Subsequently, we evaluated models including an extra variable, either B-score, mJSW, or osteophyte score, depending on the specific model. Finally, we developed a comprehensive model incorporating all DXA-derived variables and demographic factors. Goodness of fit was evaluated using AIC and BIC, while discriminative ability was assessed using AUC or C-index for logistic and Cox regression models, respectively.

To further evaluate the discriminative ability of the models, we plotted receiver operating characteristic (ROC) curves for HES-kOA and TKR at 5 years. Chi-squared tests were employed to examine the equality of the area under the curves.

Analyses were conducted using Stata version 17 (StataCorp. 2021. *Stata Statistical Software: Release 17*. College Station, TX: StataCorp LLC.) and python 3.10.5.

## Results

### Participant characteristics

In total 37,843 participants had SSM and clinical data available (Figure 1). A total of 358 participants underwent TKR (1.0%; Table 1), with an average time to surgery of 2.1 years (SD=1.4 years; males= 2.2 years [1.5], females= 2.0 years [1.3]). There were 538 deaths during the study period. The sub-sample with osteophyte data (n=6,719) were comparable with respect to their baseline demographics (Table 1), but the proportion of individuals with TKR was slightly higher (1.6%, n=110). The average time to TKR in this cohort was 2.6 years [SD: 1.5; males: 3.0 [1.5], females: 2.3 [1.4]) and there were 93 deaths.

**Figure 1:**
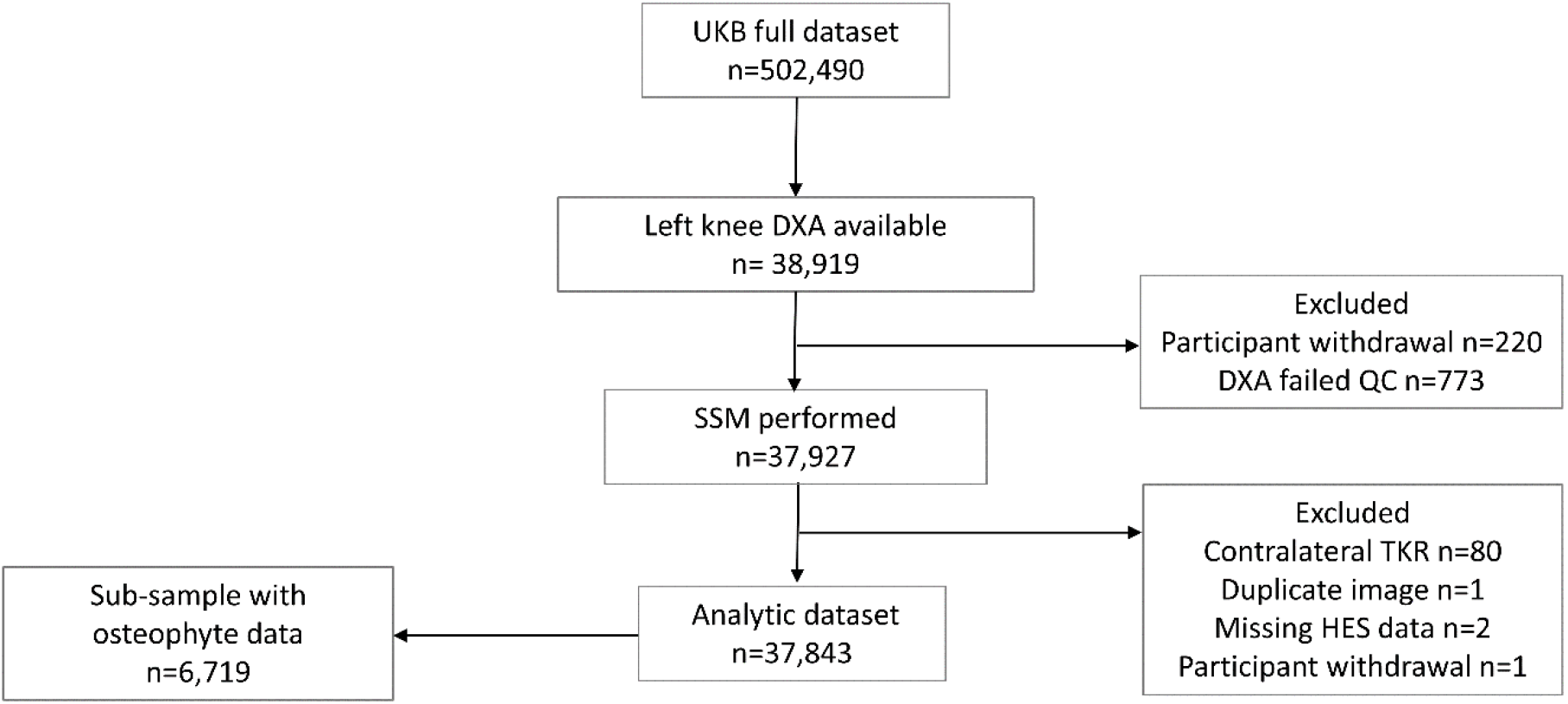
Flow Diagram of Participant Progression through the Study. At the time of the analysis, approximately 39,000 left knee DXA scans were available. DXA images underwent a comprehensive assessment to determine their suitability for inclusion in the SSM. Reasons for exclusion included: poor image quality, artefacts, positioning issues, short femoral or tibial shafts, and search failure. A total of 220 participants withdrew from the study, and an additional 80 participants were excluded due to having undergone TKR on the contralateral knee before obtaining the DXA image of the left knee. All participants in the analytic dataset had SSM data available, which was used to derive B-score and minimum joint space width (mJSW). Within this dataset, a sub-sample of 6,719 participants had additional osteophyte data available. This sub-sample, comprising participants with B-scores, mJSW and osteophyte data, were used to develop an imaging biomarker for predicting TKR. Abbreviations: QC, quality control; SSM, statistical shape model; TKR, total knee replacement; UKB, UK Biobank.

**Table 1:** Descriptive characteristics of the total study population and the subset in which osteophytes were assessed. The table presents the demographic and clinical characteristics of the study population, comprising all participants with statistical shape modelling data. The left-hand side provides information for the full dataset, while the right-hand side focuses on the sub-sample of participants who had additional osteophyte data available. Abbreviations: HES-kOA, hospital-diagnosed knee osteoarthritis; mJSW, minimum joint space width; SD, standard deviation; TKR, total knee replacement.

|  | Full dataset |  |  | Osteophyte subset |  |  |
| --- | --- | --- | --- | --- | --- | --- |
|  | Female | Male | Total | Female | Male | Total |
|  | N=19710 | N=18133 | N=37844 | N=3403 | N=3316 | N=6719 |
|  | *Mean (SD)* |  |  | *Mean (SD)* |  |  |
| Age (years) | 63.03 (7.41) | 64.41 (7.64) | 63.69 (7.55) | 62.08 (7.25) | 63.48 (7.65) | 62.77 (7.48) |
| Height (cm) | 163.64 (6.41) | 177.23 (6.62) | 170.15 (9.40) | 163.36 (6.43) | 176.90 (6.63) | 170.04 (9.40) |
| Weight (kg) | 68.11 (12.91) | 83.08 (13.38) | 75.29 (15.12) | 68.79 (12.70) | 83.65 (13.71) | 76.12 (15.15) |
| mJSW medial compartment (mm) | 3.63 (0.60) | 4.28 (0.71) | 3.94 (0.73) | 3.55 (0.70) | 4.24 (0.81) | 3.89 (0.83) |
| mJSW lateral compartment (mm) | 3.78 (0.83) | 4.65 (0.84) | 4.20 (0.94) | 3.97 (0.94) | 4.82 (0.94) | 4.39 (1.03) |
| B-score TKR | -0.10 (0.99) | 0.13 (1.02) | 0.01 (1.01) | -0.04 (1.02) | 0.08 (1.01) | 0.02 (1.01) |
| B-score HES-kOA | -0.14 (1.00) | 0.21 (1.01) | 0.03 (1.02) | -0.05 (1.02) | 0.13 (1.02) | 0.04 (1.02) |
|  | *N (%)* |  |  | *N (%)* |  |  |
| HES-kOA |  |  |  |  |  |  |
| no | 19027 (96.53%) | 17335 (95.60%) | 36362 (96.08%) | 3230 (94.92%) | 3144 (94.81%) | 6374 (94.87%) |
| yes | 684 (3.47%) | 798 (4.40%) | 1482 (3.92%) | 173 (5.08%) | 172 (5.19%) | 345 (5.13%) |
| TKR |  |  |  |  |  |  |
| no | 19526 (99.06%) | 17960 (99.05%) | 37486 (99.05%) | 3341 (98.18%) | 3268 (98.55%) | 6609 (98.36%) |
| yes | 185 (0.94%) | 173 (0.95%) | 358 (0.95%) | 62 (1.82%) | 48 (1.45%) | 110 (1.64%) |
| Osteophyte grade |  |  |  |  |  |  |
| grade 0-1 |  |  |  | 2423 (71.20%) | 2456 (74.07%) | 4879 (72.61%) |
| grade 2-3 |  |  |  | 980 (28.80%) | 860 (25.93%) | 1840 (27.39%) |

### Relationship between knee shape and kOA outcomes

We initially evaluated individual modes of variation (KSMs) in relation to kOA outcomes. Adjusted analyses results are presented in Table 2, while unadjusted associations can be found in Supplementary Table 5. A description of each KSM is provided in Supplementary Table 6.

**Table 2:** Associations between the top 10 knee shape modes and knee osteoarthritis outcomes. Hazard ratios (HRs) and odds ratios (ORs) indicate the change in risk of total knee replacement (TKR) and hospital diagnosed knee osteoarthritis (HES-kOA) per standard deviation increase in knee shape mode (KSM). Models are adjusted for age, sex, height and weight. 95% confidence intervals (CI) are provided. Associations that met the Bonferroni-significant threshold of *p*≤0.005 are shown in bold.

| TKR |  |  |  |  |
| --- | --- | --- | --- | --- |
| KSM | HR | 95% CI |  | p-value |
| 1 | 0.89 | 0.81 | 0.99 | 0.03 |
| 2 | 1.12 | 1.01 | 1.24 | 0.04 |
| 3 | 1.06 | 0.95 | 1.19 | 0.28 |
| 4 | 0.87 | 0.78 | 0.97 | 0.01 |
| 5 | 1.08 | 0.97 | 1.19 | 0.16 |
| 6 | 0.87 | 0.78 | 0.97 | 0.01 |
| 7 | 1.45 | 1.31 | 1.62 | **4.47E-12** |
| 8 | 1.69 | 1.52 | 1.88 | **1.01E-22** |
| 9 | 0.81 | 0.73 | 0.91 | **2.69E-04** |
| 10 | 0.98 | 0.88 | 1.09 | 0.69 |
| HES-kOA |  |  |  |  |
| KSM | OR | 95% CI |  | p-value |
| 1 | 0.92 | 0.87 | 0.97 | **1.61E-03** |
| 2 | 1.04 | 0.98 | 1.10 | 0.16 |
| 3 | 0.97 | 0.91 | 1.02 | 0.25 |
| 4 | 1.00 | 0.95 | 1.06 | 0.96 |
| 5 | 0.99 | 0.94 | 1.04 | 0.75 |
| 6 | 0.95 | 0.90 | 1.00 | 0.06 |
| 7 | 1.26 | 1.19 | 1.33 | **2.76E-17** |
| 8 | 1.32 | 1.25 | 1.40 | **1.02E-24** |
| 9 | 0.90 | 0.85 | 0.95 | **2.65E-04** |
| 10 | 0.92 | 0.87 | 0.97 | **2.89E-03** |

KSMs 7, 8, and 9 showed strong evidence of an association with TKR. Specifically, for each standard deviation (SD) increase in KSM9 there was a 19% reduced risk of TKR, while analogous increases in KSM7 and KSM8 were associated with a 45% and 69% increased risk, respectively.

Five KSMs displayed strong statistical evidence of an association with HES-kOA. Increases in modes 1, 9, and 10 were linked to reduced risks of 8%, 10%, and 8%, whilst increases in modes 7 and 8 were associated with a 26% and 32% increased risk, respectively.

Given that variations in knee shape could be spread over multiple modes, we next assessed the relationship between B-score, a single variable combining shape information across all KSMs, and kOA outcomes. B-score distribution is shown in Supplementary Figure 3.

We observed strong positive associations between B-scores and our outcomes (Figure 2; additional data in Supplementary Table 7). After adjustment, each SD increase in B-score was associated with a 2.3-fold higher risk of TKR (HR=2.32 [2.13, 2.54]). Similar results were found in the sex-stratified analysis (Supplementary Table 7). Additionally, each SD increase in B-score was associated with increased odds of HES-kOA, with an OR of 1.80 (1.71, 1.89). Stratifying by sex did not alter these findings.

**Figure 2:**
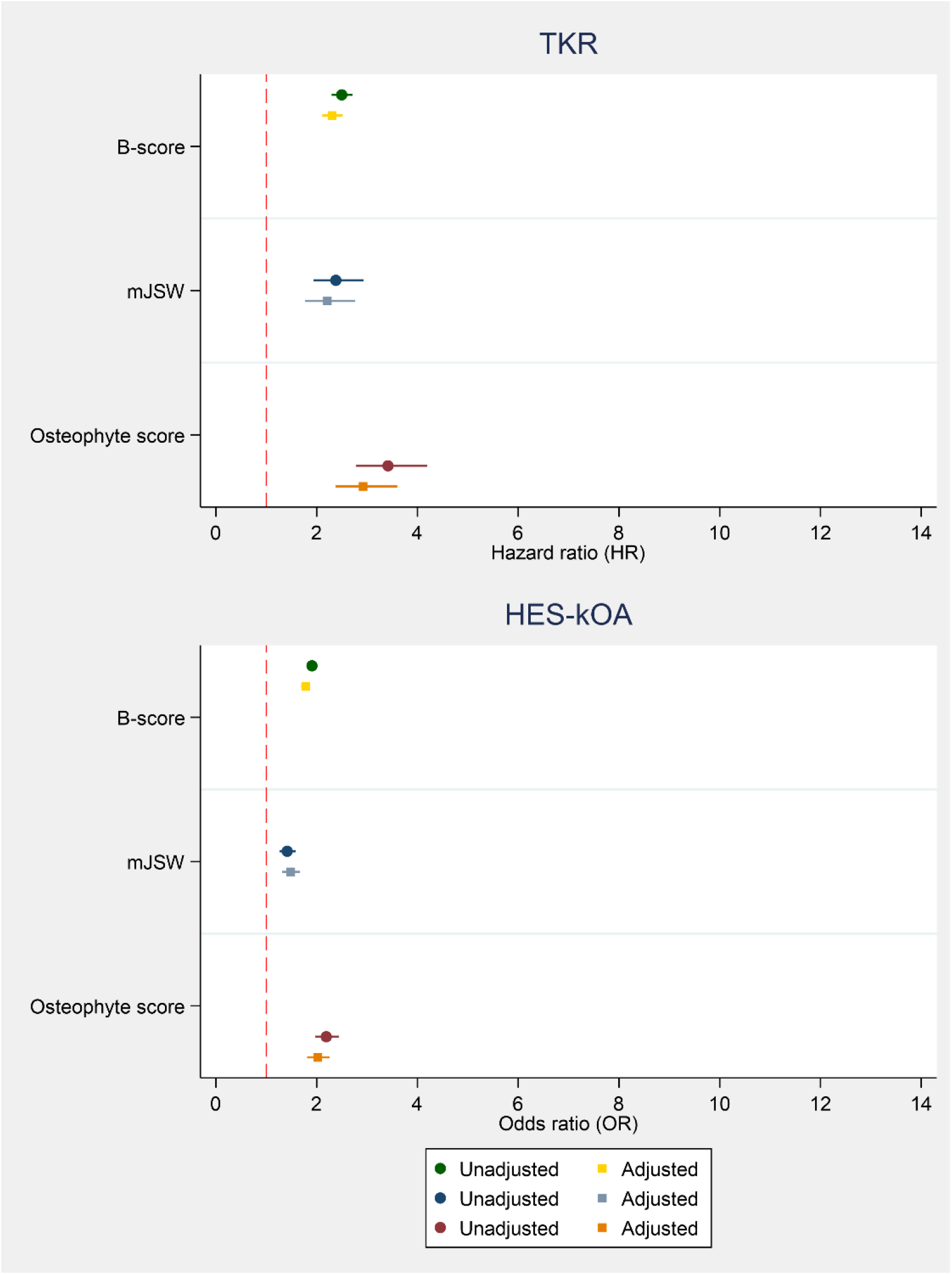
Association of B-scores, osteophyte score and mJSW with TKR and HES-kOA. The associations of B-score, mJSW, and osteophyte score with total knee replacement (TKR) are presented in the top panel, while the corresponding associations with hospital diagnosed knee osteoarthritis (HES-kOA) are displayed in the bottom panel. For B-score, the hazard ratios (HRs) and odds ratios (ORs) quantify the difference in risk associated with a standard deviation (SD) increase in the score. Regarding mJSW, the HRs and ORs represent the change in risk for individuals in the second, third, or fourth quartile of mJSW in the medial compartment, compared with those in the first quartile. For osteophyte score, (based on the sum of osteophyte grades), the HRs and ORs represent the difference in risk per unit increase in score.

Figure 3 depicts joint shape variations associated with B-scores ±2SD from the healthy group mean. An increase in B-score is linked with increasing varus alignment accompanied by reduced medial joint space width, widening of the femoral articular surface, lateral patellar displacement, and a potential upward displacement of the fibular head.

**Figure 3:**
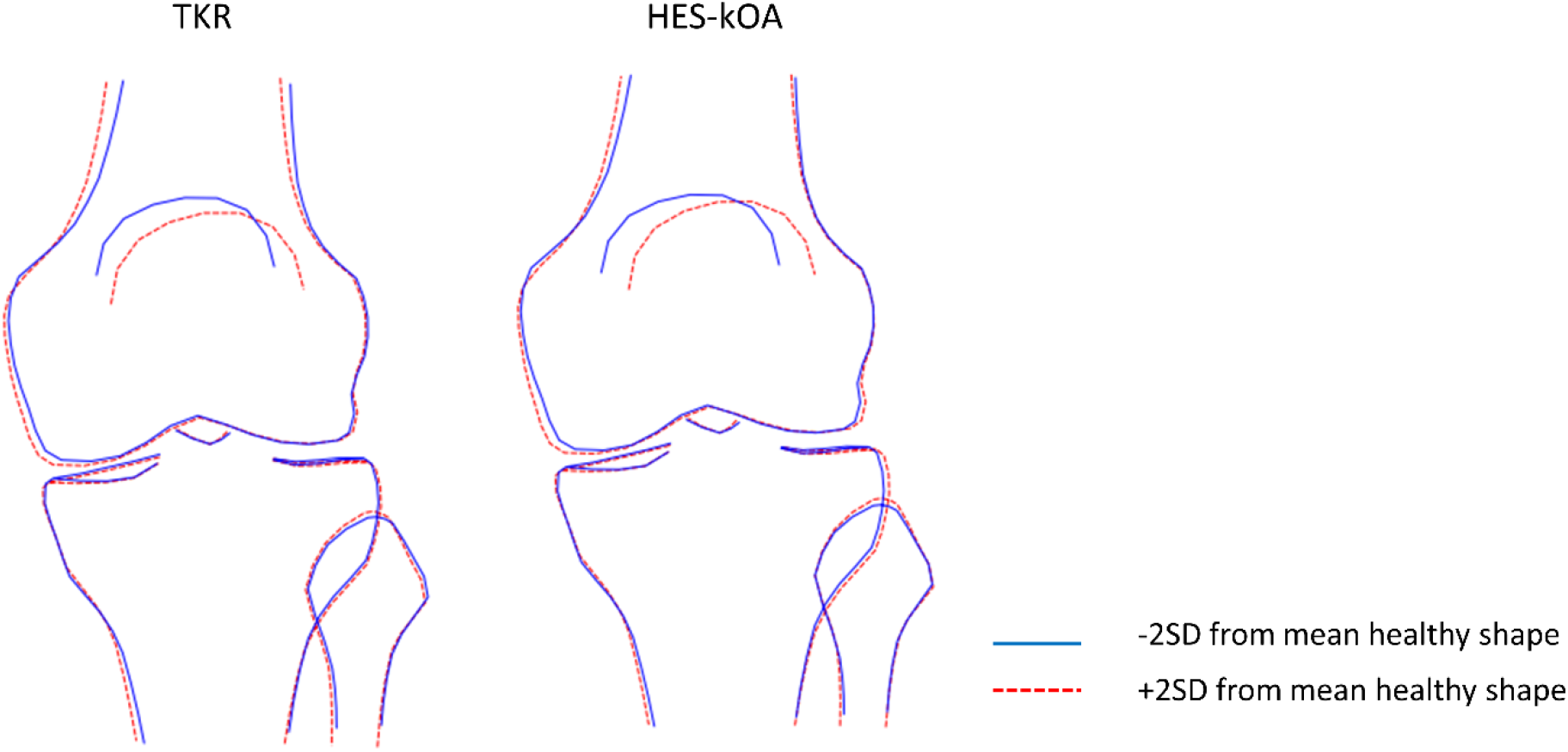
Examples of differences in bone shape corresponding to an increase and decrease in B-scores. B-scores were obtained by projecting all statistical knee shape modes (KSMs) onto a vector connecting healthy and diseased knee joint shapes. The diseased population included individuals who underwent TKR (left) or had hospital-diagnosed knee osteoarthritis (HES-kOA) (right). The figure illustrates shape changes associated with ±2 standard deviations (SD) from the mean B-score. The solid blue line depicts the shape at −2SD, while the dashed red line represents the shape at +2SD.

### Relationship between mJSW and kOA outcomes

Results are presented in Supplementary Table 8. Individuals in the first quartile of medial mJSW, representing the narrowest mJSW, had a 2.4-fold increased risk of TKR and a 54% higher risk of HES-kOA, compared with those in the fourth quartile (HR for TKR = 2.44 [1.77 to 3.38]; OR for HES-kOA = 1.54 [1.31 to 1.81]). Conversely, when examining lateral mJSW, the first and second quartiles showed a lower risk of TKR when compared with the fourth.

Considering the binary mJSW variable, individuals in the first quartile on the medial side had a 2.2-fold higher risk of TKR (HR=2.21 [1.76, 2.76]), compared with individuals in higher quartiles. They also experienced a 48% increased risk of HES-kOA (OR=1.48 [1.31, 1.67]), as depicted in Figure 2.

### Relationship between osteophytes and kOA outcomes

Supplementary Table 9 displays the prevalence of manually graded osteophytes(n=6,719). The regression analysis (Table 3, with unadjusted results in Supplementary Table 10) revealed a consistent pattern of increasing HRs and ORs for kOA outcomes with higher osteophyte grades. This was apparent across all anatomical sites.

**Table 3:** Association of osteophyte grades with TKR and HES-kOA (n=6,719). For individual osteophyte grades, hazard ratios (HRs) and odds ratios (ORs) represent the change in risk compared with grade 0 osteophytes. Models were adjusted for age, sex, height and weight. CI, 95% confidence intervals; HES-kOA, hospital diagnosed knee osteoarthritis; TKR, total knee replacement.

|  | TKR |  |  |  | HES-kOA |  |  |  |
| --- | --- | --- | --- | --- | --- | --- | --- | --- |
|  | HR | 95% CI |  | *p*-value | OR | 95% CI |  | *p*-value |
| **Medial femur** |  |  |  |  |  |  |  |  |
| grade 1 | 2.00 | 1.22 | 3.29 | 0.01 | 1.43 | 1.09 | 1.87 | 0.01 |
| grade 2 | 8.79 | 5.21 | 14.82 | $3.35 \times 10^{-16}$ | 5.93 | 4.22 | 8.34 | $1.24 \times 10^{-24}$ |
| grade 3 | 17.55 | 10.05 | 30.65 | $7.04 \times 10^{-24}$ | 8.27 | 5.23 | 13.10 | $2.04 \times 10^{-19}$ |
| **Lateral femur** |  |  |  |  |  |  |  |  |
| grade 1 | 2.35 | 1.21 | 4.60 | 0.01 | 2.00 | 1.32 | 3.05 | $1.24 \times 10^{-24}$ |
| grade 2 | 9.56 | 5.61 | 16.28 | $1.01 \times 10^{-16}$ | 7.13 | 4.74 | 10.74 | $2.04 \times 10^{-19}$ |
| grade 3 | 11.81 | 6.48 | 21.54 | $8.09 \times 10^{-16}$ | 5.44 | 3.09 | 9.57 | $4.12 \times 10^{-09}$ |
| **Medial tibia** |  |  |  |  |  |  |  |  |
| grade 1 | 3.45 | 2.24 | 5.31 | $1.81 \times 10^{-08}$ | 2.84 | 2.24 | 3.60 | $8.65 \times 10^{-18}$ |
| grade 2 | 9.82 | 5.44 | 17.72 | $3.35 \times 10^{-14}$ | 5.42 | 3.47 | 8.45 | $9.67 \times 10^{-14}$ |
| grade 3 | 22.12 | 10.04 | 48.74 | $1.55 \times 10^{-14}$ | 14.52 | 6.66 | 31.64 | $1.71 \times 10^{-11}$ |
| **Lateral tibia** |  |  |  |  |  |  |  |  |
| grade 1 | 2.82 | 1.82 | 4.37 | $3.17 \times 10^{-06}$ | 2.01 | 1.59 | 2.54 | $6.47 \times 10^{-09}$ |
| grade 2 | 7.66 | 3.92 | 14.98 | $2.66 \times 10^{-09}$ | 5.81 | 3.71 | 9.09 | $1.38 \times 10^{-14}$ |
| grade 3 | 33.22 | 16.64 | 66.31 | $3.01 \times 10^{-23}$ | 14.53 | 7.22 | 29.21 | $5.97 \times 10^{-14}$ |

An osteophyte score of 3 (based on the sum of the osteophyte grades) was associated with an 18-fold increased risk of TKR (HR=18.30 [9.98, 33.57]), and a 9-fold increased risk of HES-kOA (OR=9.10 [6.54, 12.67]) compared to a score of 0 (Supplementary Table 11). Figure 2 visually depicts the incremental impact per unit increase in the osteophyte score.

### Performance of multivariable models in predicting TKR

Table 4 displays the predictive performance of different models for TKR, combining demographic variables (age, sex, height and weight), B-score, binary mJSW and osteophyte score in the sub-group of 6,719 participants.

**Table 4:** Predictive performance of B-scores, binary mJSW and osteophyte score with TKR and HES-kOA (n=6,719). The table includes results of univariable and multivariable models (left), and measures of model fit and discrimination (right). Demographic characteristics include age, sex, height and weight. For B-score, HRs and ORs indicate the change in risk per standard deviation increase in B-score. For mJSW, HRs and ORs reflect the risk difference between the first quartile (on the medial compartment) compared with quartiles two, three, and four. Osteophyte score was entered into the model as a linear term and therefore HRs and ORs represent the difference in risk per unit increase in score. When evaluating the predictive accuracy of the time-to-event (cox) models we used Harrell’s C-index. For logistic regression models, where the outcome was binary, we used the AUC. The most parsimonious model, containing the predictors B-score, osteophyte grade and demographic variables, is shown in bold. Abbreviations: AIC, Akaike information criterion; AUC, area under the receiver operating characteristic curve; BIC, Bayesian information criterion; CI, 95 % confidence intervals; C-index, Harrell’s concordance index; Demo, demographic variables; OP, osteophyte score; HR, hazard ratio; mJSW, minimum joint space width; OR, odds ratio; TKR, total knee replacement.

| TKR |  |  |  |  |  |  |  |
| --- | --- | --- | --- | --- | --- | --- | --- |
| model | HR | 95% CI |  | p-value | AIC | BIC | C-stat |
| B-score | 2.67 | 2.25 | 3.17 | $2.86 \times 10^{-29}$ | 1808.50 | 1815.31 | 0.75 |
| B-score + demo | 2.40 | 2.00 | 2.87 | $9.47 \times 10^{-22}$ | 1757.53 | 1791.59 | 0.81 |
| B-score + demo + mJSW | 2.29 | 1.89 | 2.76 | $8.84 \times 10^{-18}$ | 1757.45 | 1798.33 | 0.81 |
| B-score + demo + OP | 1.80 | 1.50 | 2.15 | $2.70 \times 10^{-10}$ | **1687.85** | **1728.72** | **0.87** |
| B-score + demo + mJSW + OP | 1.72 | 1.42 | 2.09 | $2.93 \times 10^{-08}$ | 1688.39 | 1736.08 | 0.87 |
| mJSW | 2.81 | 1.94 | 4.09 | $5.98 \times 10^{-08}$ | 1898.62 | 1905.44 | 0.62 |
| mJSW + demos | 2.29 | 1.54 | 3.41 | $4.50 \times 10^{-05}$ | 1829.75 | 1863.81 | 0.77 |
| mJSW + demos + B-score | 1.36 | 0.90 | 2.05 | 0.15 | 1757.45 | 1798.33 | 0.81 |
| mJSW + demos + OP | 1.91 | 1.29 | 2.82 | $1.13 \times 10^{-03}$ | 1718.01 | 1758.89 | 0.86 |
| mJSW + demo + B-score + OP | 1.29 | 0.85 | 1.96 | 0.23 | 1688.39 | 1736.08 | 0.87 |
| OP | 3.42 | 2.78 | 4.19 | $8.16 \times 10^{-32}$ | 1761.45 | 1768.26 | 0.80 |
| OP + demo | 2.92 | 2.37 | 3.60 | $8.54 \times 10^{-24}$ | 1726.42 | 1760.48 | 0.85 |
| OP + demo + B-score | 2.35 | 1.90 | 2.90 | $3.33 \times 10^{-15}$ | **1687.85** | **1728.72** | **0.87** |
| OP + demo + mJSW | 2.82 | 2.30 | 3.48 | $9.26 \times 10^{-23}$ | 1718.01 | 1758.89 | 0.86 |
| OP + demo + B-score + mJSW | 2.34 | 1.89 | 2.89 | $4.19 \times 10^{-15}$ | 1688.39 | 1736.08 | 0.87 |
| HES-KOA |  |  |  |  |  |  |  |
| model | OR | 95% CI |  | p-value | AIC | BIC | AUC |
| B-score | 1.98 | 1.78 | 2.20 | $1.73 \times 10^{-36}$ | 2564.08 | 2577.70 | 0.68 |
| B-score + demo | 1.85 | 1.66 | 2.07 | $6.99 \times 10^{-28}$ | 2527.02 | 2567.90 | 0.71 |
| B-score + demo + mJSW | 1.82 | 1.63 | 2.04 | $2.63 \times 10^{-25}$ | 2527.82 | 2575.51 | 0.71 |
| B-score + demo + OP | 1.59 | 1.42 | 1.77 | $4.84 \times 10^{-16}$ | **2422.33** | **2470.02** | **0.75** |
| B-score + demo + mJSW + OP | 1.57 | 1.40 | 1.76 | $1.98 \times 10^{-14}$ | 2423.51 | 2478.01 | 0.75 |
| mJSW | 1.65 | 1.31 | 2.07 | $1.97 \times 10^{-05}$ | 2707.38 | 2721.01 | 0.55 |
| mJSW + demo | 1.55 | 1.21 | 1.98 | $4.61 \times 10^{-04}$ | 2636.44 | 2677.31 | 0.65 |
| mJSW + demo + B-score | 1.15 | 0.89 | 1.48 | 0.27 | 2527.82 | 2575.51 | 0.71 |
| mJSW + demo + OP | 1.44 | 1.13 | 1.84 | $3.78 \times 10^{-03}$ | 2481.69 | 2529.38 | 0.73 |
| mJSW + demo + B-score + OP | 1.13 | 0.87 | 1.46 | 0.36 | 2423.51 | 2478.01 | 0.75 |
| OP | 2.19 | 1.97 | 2.44 | 0.00E+00 | 2513.93 | 2527.56 | 0.70 |
| OP + demo | 2.02 | 1.81 | 2.26 | $1.33 \times 10^{-35}$ | 2487.86 | 2528.73 | 0.73 |
| OP + demo + B-score | 1.80 | 1.60 | 2.01 | $2.70 \times 10^{-24}$ | **2422.33** | **2470.02** | **0.75** |
| OP + demo + mJSW | 2.00 | 1.79 | 2.24 | $7.83 \times 10^{-35}$ | 2481.69 | 2529.38 | 0.73 |
| OP + demo + B-score + mJSW | 1.79 | 1.60 | 2.01 | $3.18 \times 10^{-24}$ | 2423.51 | 2478.01 | 0.75 |

When assessing the B-score’s relationship with TKR, the univariable model yielded a HR of 2.67 (2.25, 3.17), which decreased to 2.40 (2.00, 2.87) upon adjustment for demographic variables. Adding osteophyte score had a greater impact compared to mJSW, resulting in HRs of 1.80 (1.50, 2.15) and 2.29 (1.89, 2.76) respectively. Combining mJSW, osteophyte score, B-score, and demographics resulted in a HR of 1.72 (1.42, 2.09).

The unadjusted HR for the mJSW-TKR association was 2.81 (1.94, 4.09), which decreased to 2.29 (1.54, 3.41) after incorporating demographic variables. Adding B-score to the latter model had a more pronounced effect compared to osteophyte score, with HRs of 1.36 (0.90, 2.05) and 1.91 (1.29, 2.82), respectively. The addition of both B-score and osteophyte score yielded an HR of 1.29 (0.85, 1.96).

In the context of the osteophyte-TKR association, the unadjusted hazard ratio (HR) was 3.42 (2.78, 4.19) per unit increase, decreasing to 2.92 (2.37, 3.60) after adjusting for demographics. Adding mJSW further reduced the HR to 2.82 (2.30, 3.48), while the inclusion of B-score had a more substantial impact, resulting in a HR of 2.35 (1.90, 2.90). Similar effects were observed in the model including both B-score and mJSW, alongside demographic variables (HR = 2.34 [1.89, 2.89]).

The model that exhibited the optimal fit, as indicated by the lowest AIC and BIC, and the highest discrimination as reflected by C-index, comprised B-score, osteophyte score and demographics.

We subsequently examined the predictive ability of this model in classifying TKR at five years. A model including demographic factors alone achieved an AUC of 0.73 (Figure 4), while the optimal model (including B-score, osteophyte score and demographics) had an AUC of 0.87 (p < 0.05).

**Figure 4:**
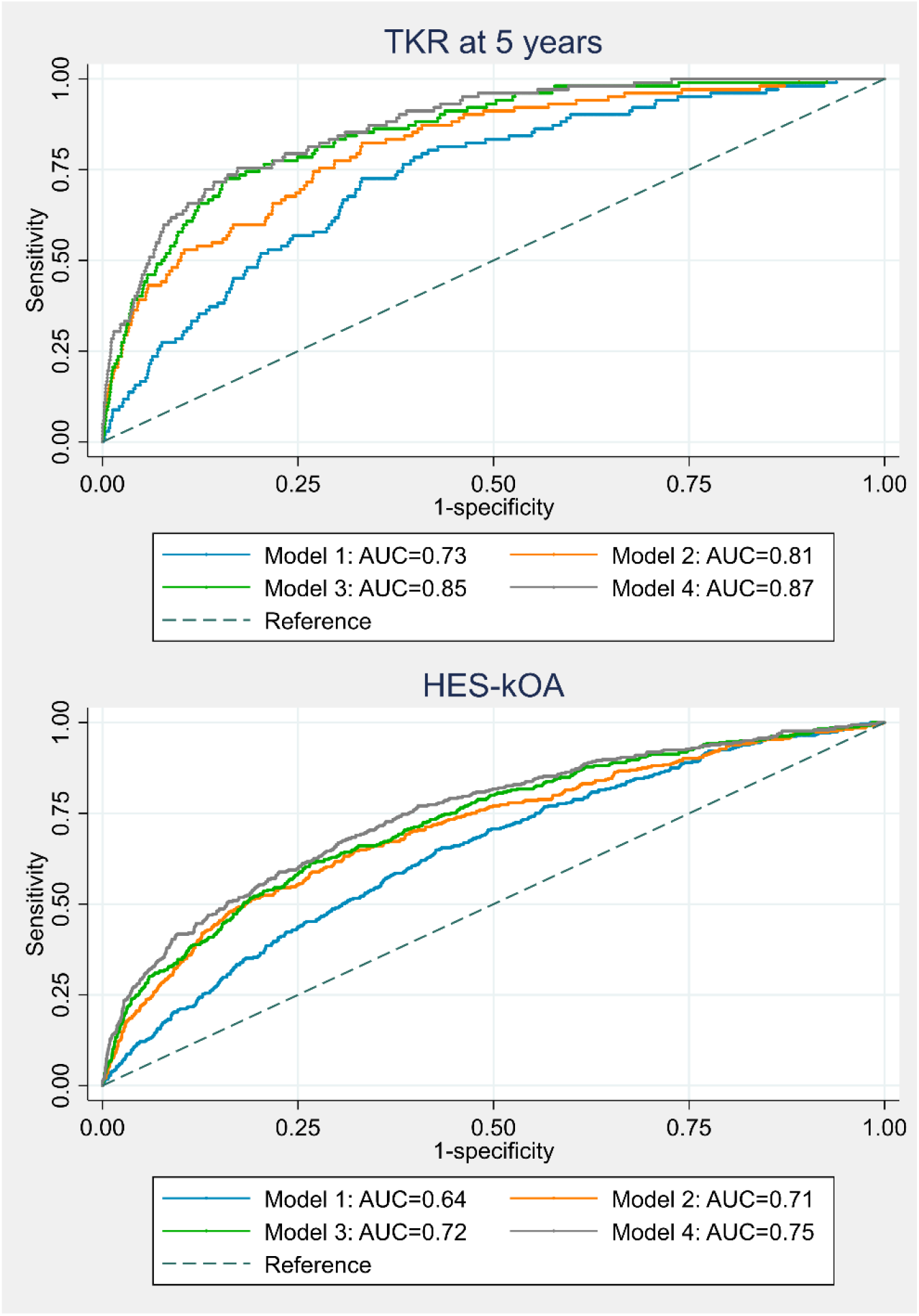
Receiver Operating Characteristic (ROC) Curve for Prediction of HES-kOA and TKR at 5 Years (n=6,719). Model 1: age, sex, height, and weight; Model 2: age, sex, height, weight, B-score; Model 3: age, sex, height, weight, binary osteophyte grade; Model 4: age, sex, height, weight, B-score, binary osteophyte grade. Abbreviation: AUC, area under the receiver operating characteristic curve.

### Performance of multivariable models in predicting HES-kOA

The associations between DXA-derived variables and HES-kOA were generally observed to be weaker when compared with those with TKR (Table 4).

The B-score-HES-kOA association partially attenuated when demographic variables were included in the model, with the OR decreasing from 1.98 (1.78, 2.20) to 1.85 (1.66, 2.07). Additional adjustment for mJSW had minimal impact (OR=1.82 [1.63, 2.04]), but incorporating osteophyte score further reduced the association (OR=1.59 [1.42, 1.77]). Including both mJSW and osteophyte score in the model resulted in a comparable OR to the model that adjusted for osteophyte score and demographics (OR=1.57 [1.40, 1.76]).

When considering mJSW, adjusting for demographic variables reduced the strength of the association, causing the OR to shift from 1.65 (95% CI: 1.31, 2.07) to 1.55 (95% CI: 1.21, 1.98). Further adjustment for B-score led to a more pronounced attenuation, yielding an odds ratio (OR) of 1.15 (95% CI: 0.89, 1.48), in contrast to the adjustment for osteophyte score which produced an OR of 1.44 (95% CI: 1.13, 1.84). Including both B-score and osteophyte score produced an OR similar to the model adjusting for demographics and B-score alone (OR = 1.13 [0.87, 1.46]).

Osteophyte score exhibited the strongest association with HES-kOA, with an OR of 2.19 (1.97, 2.44) in univariable analyses and 2.02 (1.81, 2.26) after adjusting for demographics. Adding mJSW as an additional adjustment did not change the estimate (OR=2.00 [1.79, 2.24]), but partial attenuation occurred when B-score was added (OR=1.80 [1.60, 2.01]). The model with B-score, mJSW, and demographics was comparable to the one with B-score and demographics (OR=1.79 [1.60, 2.01]).

As seen for TKR, the model that exhibited the optimal fit and discrimination comprised the B-score, osteophyte score and demographic variables. When considering demographic variables alone, the model yielded an AUC of 0.64. However, when the B-score and osteophyte score were incorporated into the model, the AUC improved to 0.75.

## Discussion

The purpose of this study was to develop an imaging biomarker derived from knee DXA scans which incorporates joint shape and to investigate its potential prognostic value in predicting TKR. In addition to capturing conventional features of radiographic kOA, namely JSN (as reflected by mJSW) and osteophytes, an automated model was developed to extract knee shape information using SSM. This information was quantified as a B-score. We found strong correlations between all of the DXA-derived features investigated and subsequent risk of TKR. Similar, although comparatively weaker associations were observed with HES-kOA. Furthermore, when we combined B-score with osteophyte score and demographic factors, we observed a significant improvement in the accuracy of predicting TKR at 5 years, compared with demographic risk factors alone (AUC=0.87 compared with 0.73).

Few studies have examined knee shape using DXA-based SSM. One small study explored knee shape changes in 109 individuals over 12 months. It found significant alterations in a particular shape mode reflecting reduced medial mJSW in kOA cases, which aligns with our findings (10). However, this investigation did not annotate osteophytes separately or observe overall shape changes like varus alignment. In contrast, our analysis revealed a strong relationship between DXA-derived osteophytes and subsequent joint replacement, consistent with our previous hip DXA study (16). Our findings also align with an earlier radiographic hip investigation which found that incorporating an SSM-derived Shape-Score into a model alongside demographic factors, clinical assessment, and radiologists scores enhanced the prediction of hip OA (27). The initial prediction model had an AUC of 0.795, which improved to 0.864 with the inclusion of the Shape-Score.

The observed trend toward varus alignment in individuals with HES-kOA and subsequent TKR in our study is consistent with the recognised varus malalignment characteristic in kOA (1, 2). This malignment may be a predisposing factor, focusing weight-bearing forces through the medial compartment causing considerable biomechanical stress. Alternatively, varus may develop due to medial JSN, a recognized feature of primary kOA. Alongside the varus alignment and reduced mJSW, there appeared to be an enlargement of the femoral articular surface. These findings coincide with previous studies using MRI-based SSM approaches, which have reported femoral condyle widening and flattening in osteoarthritic knees (13, 28–30).

By employing a B-score, we were able to examine independent associations between knee shape and other features related to kOA, namely mJSW and osteophytes. Whereas B-score, osteophytes and mJSW were all related to TKR and HES-kOA in univariable analyses, the association between mJSW and these outcomes attenuated following inclusion of B-score, suggesting that mJSW is encompassed by the SSM and the resulting B-score. In contrast, B-score and osteophyte score were both retained as independent predictors of clinical outcomes, and their combination increased model performance. That said, the association between osteophytes and risk of TKR/HES-kOA was partially attenuated by inclusion of B-score. Since our SSM template excluded osteophytes, these are unlikely to have directly contributed to the B-score, suggesting that joint malalignment may contribute to the relationship between osteophytes and kOA progression, as previously proposed (31). Demographic factors such as age, sex, and weight also strongly predicted risk of TKR/HES-kOA, implying imaging biomarkers need to be considered as complementing these factors in patient assessment rather than replacing them.

The exclusion of participants with previous TKR allowed us to assess the predictive capability of DXA-derived imaging biomarkers for future TKR. On the other hand, participants could have had pre-existing HES-kOA at the time of their DXA scan. It was reassuring that broadly consistent relationships were seen between imaging biomarkers and HES-kOA and TKR. Nonetheless, given we were only able to examine cross-sectional relationships with HES-kOA, further studies are necessary to determine whether DXA biomarkers are also useful in predicting subsequent HES-kOA when obtained in those with early disease.

Several approaches have been used to extract additional features on knee images for OA diagnosis and prognosis, including Deep Learning methods, as has previously been applied to knee radiographs (32–34). Our investigation is novel in exploring whether an imaging biomarker incorporating knee shape might be useful in predicting TKR when applied to knee DXA scans. This was made possible by application of SSM to DXA images obtained from a large sample of individuals from UKB, in whom follow-up data for THR via HES linkage was available. Given the study’s scale, our SSM could serve as a reference model, making replication, validation and clinical application more feasible. Indeed, the SSM, along with the BoneFinder search model, will be made publicly available on the BoneFinder website. Another strength of our study is that, unlike the majority of previous studies using SSM to characterise knee shape, which generally focused on the tibia and femur, our SSM also incorporated the superior patella and fibula.

A limitation of our study is that whereas knee shape data were obtained in all those with available DXA scans, osteophytes, which required manual annotation, were only obtained in a subset. Nevertheless, this smaller sample had sufficient numbers of individuals with TKR to compare the predictive value of different models. A further limitation is that HES linkage may have missed some cases of TKR through failure to capture procedures performed in the private sector. Moreover, the HES data lacks laterality but this limitation is more likely to reduce effect sizes rather than introduce bias into the study. Lack of generalisability may also be an issue given UKB is 95% white and has lower rates of all-cause mortality compared with the population at large, which aligns with the well-known “healthy volunteer” effect (35). In addition, those 11% of UKB participants recruited from Scotland and Wales were excluded as they have separate systems to HES linkage. Finally, it is important to stress that the present study aimed to explore the utility of DXA-based measures of knee shape in predicting clinical outcomes related to kOA, and to evaluate whether these provide additional information compared to conventional radiographic measures, including JSW and osteophytes. Further studies, including validation in an external dataset, are required to establish the generalizability and reliability of this approach prior to clinical use.

In summary, our study highlights the potential value of SSM as a tool for characterising joint shape in knee DXA scans. Importantly, the development of a DXA-derived imaging biomarker combining knee shape and osteophyte formation shows promise in identifying individuals at high risk of progression to TKR, particularly when combined with demographic factors. Further studies are justified to examine the utility of using DXA scans, which are widely available and involve much lower doses of radiation compared with conventional X-rays, to identify individuals who are likely to benefit from interventions aimed at slowing kOA progression.

## Data Availability

The data from this study will be available from UK Biobank at a forthcoming data release. Users must be registered with UK Biobank to access their resources [https://bbams.ndph.ox.ac.uk/ams/].

## Acknowledgements

The authors would like to express their gratitude to Dr. David Wilson, Consultant Interventional Musculoskeletal Radiologist and honorary Clinical Lecturer at Aberdeen University, for his valuable contribution to the development of the DXA-based osteophyte Atlas. They also extend their appreciation to all the participants of the UK Biobank study.

## Author contributions

Each author has made significant contributions to the study’s conception, design, data acquisition, analysis, and interpretation. Furthermore, all authors helped draft the article before approving the final version of this manuscript.

