## Supplementary material for "Dual-energy X-ray absorptiometry derived knee shape may provide a useful imaging biomarker for predicting total knee replacement: findings from a study of 37,843 people in UK Biobank"

Supplementary Table 1: A list of the ICD-9 and ICD-10 revision codes used for the categorisation of HES-kOA and the OPCS-4 codes for identification of knee replacement.

| **ICD 10 code** | **Condition** |
| --- | --- |
| M17 | Gonarthrosis [arthrosis of knee]. |
| M170 | Primary gonarthrosis, bilateral. |
| M171 | Other primary gonarthrosis. |
| M179 | Gonarthrosis, unspecified. |
| M1906 | Primary arthrosis of other joints, lower leg. |
| M1996 | Arthrosis, unspecified, lower leg. |
| **ICD 9 code** | **Condition** |
| 71536 | Osteoarthrosis, localized, not specified whether primary or secondary, lower leg. |
| 71516 | Localised, primary osteoarthrosis and allied disorders, lower leg. |
| **OPSC-4 code** |  |
| W40 | Total prosthetic replacement of knee joint using cement. |
| W41 | Total prosthetic replacement of knee joint not using cement. |
| W42 | Other total prosthetic replacement of knee joint. |

Cases of hospital diagnosed knee OA (HES-kOA) and total knee replacement (TKR) were identified via linkage to the HES database, which records details of all admissions, outpatient appointments, and A&E attendances at National Health Service (NHS) hospitals in England. The HES database uses the International Classification of Diseases, 9th (ICD-9) and 10th (ICD-10) revision codes to classify diseases and health conditions, while surgical procedures are classified using the Office of Population Censuses and Surveys (OPCS) Classification of Surgical Operations and Procedures, version 4 codes (1). In this study, we obtained the relevant codes from the UKB data fields 41270 (ICD-10), 41271 (ICD-9), and 41273 (OPCS-4), adopting the codes utilized in the study conducted by Zengini et al (2).

Supplementary table 2: Results of the 3-fold cross-validation experiment used to evaluate the automated search model.

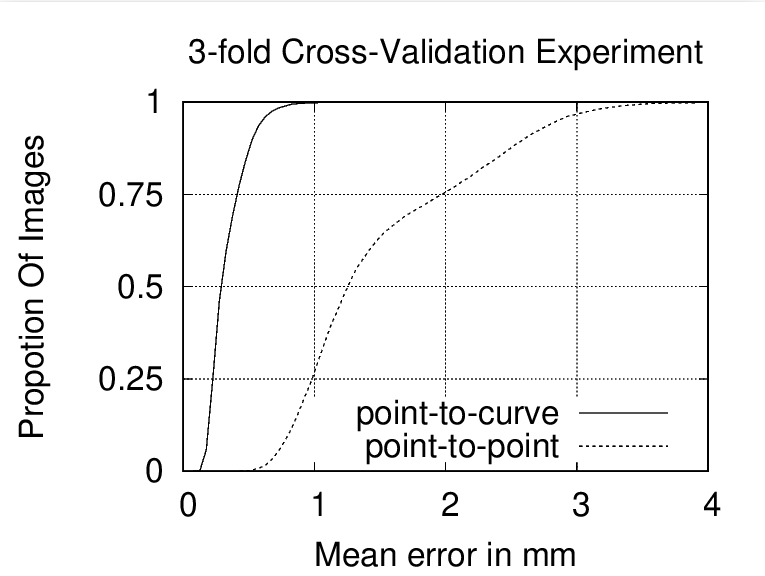

The model was evaluated using the Euclidean distance between the automatic points and the manual points (i.e., point-to-point error), and the curve passing through manual points (i.e., point-to-curve error). Mean point-to-point error, and point-to-curve error were under 3mm, and 0.7mm respectively in 95% of the images.

Supplementary Table 3: Number of images corrected for point placement during development of the SSM.

| **Bone** | **No. of images with manual point corrections** | **Average point-to-point correction distance (mm)** |
| --- | --- | --- |
| Femur | 812 | 1.8 |
| Tibia | 1,074 | 1.7 |
| Fibula | 2,253 | 4.6 |
| Patella | 1,714 | 4.9 |
| **Total** | **4,214** |  |

The automated search model placed points to 31,207 images. Manual point corrections were made to 4,214 images.

Supplementary Table 4: Creating a DXA-based osteophyte score.

| **Sum of visually graded OPs** | **n** | **OP score** | **n** |
| --- | --- | --- | --- |
| 0 | 3,122 | 0 | 3,122 |
| 1 | 1,757 | 1 | 1,757 |
| 2 | 964 | 2 | 1,393 |
| 3 | 429 |  |  |
| 4 | 169 | 3 | 447 |
| 5 | 95 |  |  |
| 6 | 62 |  |  |
| 7 | 44 |  |  |
| 8 | 32 |  |  |
| 9 | 16 |  |  |
| 10 | 18 |  |  |
| 11 | 6 |  |  |
| 12 | 5 |  |  |

Osteophytes were evaluated in 6,719 DXA images. In the table, the first two columns present the total cumulative sum of manually graded osteophytes, accompanied by the count of individual knees displaying each respective value. Using this cumulative value, an osteophyte score ranging from 0 to 3 was calculated for each knee. The final two columns of the table depict the frequency of knees falling within each of these osteophyte scores.

Supplementary Table 5: Unadjusted association of top 10 shape modes with kOA (n=37,843).

| TKR | | | | |
| --- | --- | --- | --- | --- |
| KSM | HR | 95% CI | | *p*-value |
| 1 | 0.81 | 0.73 | 0.90 | **8.15 x10^-05^** |
| 2 | 1.19 | 1.07 | 1.31 | **1.22 x10^-03^** |
| 3 | 1.03 | 0.93 | 1.14 | 0.56 |
| 4 | 0.87 | 0.79 | 0.97 | 0.01 |
| 5 | 1.11 | 1.00 | 1.23 | 0.05 |
| 6 | 0.84 | 0.75 | 0.93 | **1.03 x10^-3^** |
| 7 | 1.52 | 1.37 | 1.69 | **1.14 x10^-14^** |
| 8 | 1.72 | 1.55 | 1.91 | **1.92 x10^-24^** |
| 9 | 0.74 | 0.67 | 0.82 | **1.65 x10^-08^** |
| 10 | 0.99 | 0.89 | 1.10 | 0.82 |
| HES-kOA | | | | |
| KSM | OR | 95% CI | | *p*-value |
| 1 | 0.85 | 0.80 | 0.89 | **4.17 x10^-10^** |
| 2 | 1.11 | 1.05 | 1.17 | **7.77 x10^-05^** |
| 3 | 0.91 | 0.86 | 0.96 | **4.21 x10^-04^** |
| 4 | 1.00 | 0.95 | 1.05 | 0.88 |
| 5 | 1.02 | 0.97 | 1.08 | 0.39 |
| 6 | 0.92 | 0.88 | 0.97 | **2.96 x10^-03^** |
| 7 | 1.30 | 1.23 | 1.37 | **8.99 x10^-22^** |
| 8 | 1.36 | 1.29 | 1.44 | **5.45 x10^-31^** |
| 9 | 0.87 | 0.82 | 0.91 | **4.75E^-08^** |
| 10 | 0.95 | 0.90 | 1.00 | 0.06 |

Hazard ratios (HRs) and odds ratios (ORs) represent the change in risk of total knee replacement (TKR) and hospital diagnosed knee osteoarthritis (HES-kOA) per standard deviation increase in knee shape mode (KSM). Associations that met the Bonferroni-significant threshold of *P*≤0.005 are shown in bold. CI, 95% confidence intervals.

Supplementary Table 6: variation described by the top 10 knee shape modes (KSMs).

| Mode  (% of variation) | Shape ±2SD | Changes Associated with a -2SD decrease |
| --- | --- | --- |
| 1 (22.9%) | 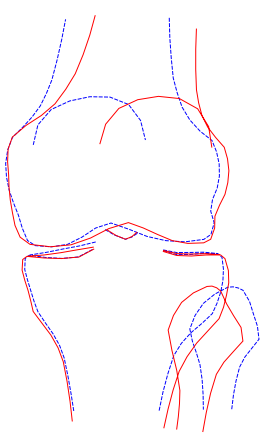 | - Varus alignment of the femur. - Medial patellar displacement. - Lateral shift of the fibula. |
| 2 (15.3%) | 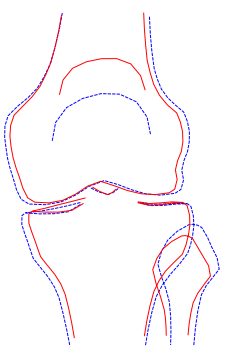 | - Lower patella height. - Widening of the Femur and Tibia. - Taller fibula head. |
| 3 (11.7%) | 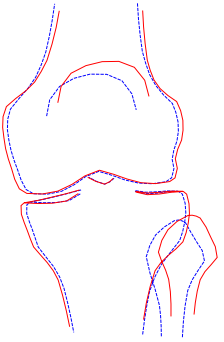 | - Lower patella height, accompanied by a medial shift. - Medial Displacement and Lowered Position of the Fibula. - Reduction in the width of the tibial plateau. - Reduction in the width of the femoral plateau and condyles. |
| 4 (7.9%) | 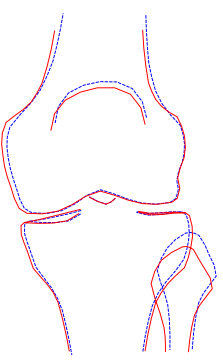 | - Elevated Position of the Fibula. - Narrowing of the Medial Femoral Width. - Slight Elevation in Patella Height. |
| 5 (5.0%) | 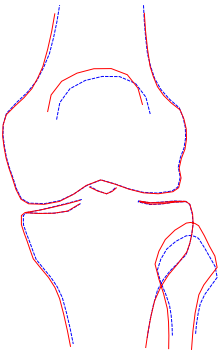 | - Moderate lateral shift and lower positioning of the patella - Downward positioning of the fibula. - Reduced depth of the fibula head. |
| 6 (4.3%) | 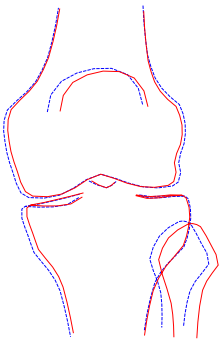 | - Moderate medial displacement of the patella. - Medial shift of the fibula. - Widening of the medial tibia. - Widening in the proximal region of the femur. |
| 7 (4.0%) | 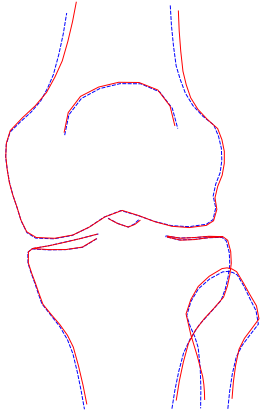 | - Medial shift in the femoral shaft. |
| 8 (3.4%) | 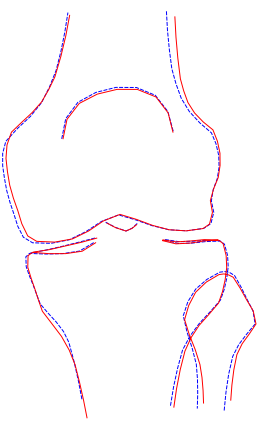 | - Narrowing of the femoral shaft, laterally. - Increased curvature beneath the medial tibial condyle. |
| 9 (3.2%) | 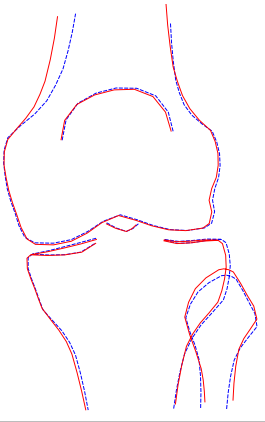 | - Narrowing of the medial femoral shaft. - Smaller fibular head. |
| 10 (2.6%) | 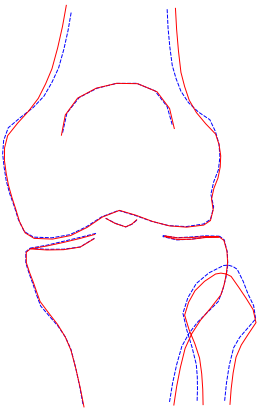 | - Narrowing of the femoral shaft. - Larger fibular head. |

+ 2SD

- 2SD

The first column in the table displays the extent of shape variation captured by each statistical knee shape mode (KSM). In the second column, visual representations of the mode are presented, depicting solid red lines for shapes corresponding to values 2 standard deviations (SD) above the mean mode shape, and dashed blue lines for shapes associated with values 2 SD below the mean mode shape. The last column offers a description of the alterations related to a 2 SD decrease in KSM.

Supplementary Table 7: Association of sex-specific and sex-combined B-scores with kOA outcomes (n=37,843).

| Unadjusted | | | | | Adjusted | | | | | |
| --- | --- | --- | --- | --- | --- | --- | --- | --- | --- | --- |
| **TKR** | | | | | | | | | | |
| **B-score** | N | HR | 95% CI | | *p*-value |  | HR | 95% CI | | *p*-value |
| Male | 18133 | 2.62 | 2.34 | 2.95 | <0.0001 |  | 2.45 | 2.17 | 2.77 | <0.0001 |
| Female | 19710 | 2.66 | 2.34 | 3.02 | <0.0001 |  | 2.36 | 2.08 | 2.69 | <0.0001 |
| Combined | 37843 | 2.52 | 2.32 | 2.74 | <0.0001 |  | 2.32 | 2.13 | 2.54 | <0.0001 |
| **HES-kOA** | | | | | | | | | | |
| **B-score** | N | OR | 95% CI | | *p*-value |  | OR | 95% CI | | *p*-value |
| Male | 18133 | 1.89 | 1.77 | 2.02 | <0.0001 |  | 1.81 | 1.69 | 1.93 | <0.0001 |
| Female | 19710 | 2.00 | 1.86 | 2.15 | <0.0001 |  | 1.82 | 1.69 | 1.96 | <0.0001 |
| Combined | 37843 | 1.93 | 1.84 | 2.03 | <0.0001 |  | 1.80 | 1.71 | 1.89 | <0.0001 |

Hazard ratios (HRs) and Odds ratios (ORs) reflect the change in risk for total knee replacement (TKR) (top panel) and hospital diagnosed knee osteoarthritis (HES-kOA) (bottom panel) per standard deviation increase in B-score. Adjusted models incorporate age, sex, height, and weight as covariates. CI, 95% confidence intervals. *p-*values akin to 0.00E+00.

Supplementary Table 8: Associations of minimum joint space width (mJSW) with HES-kOA and TKR (n=37,843).

|  | Unadjusted | | | |  | Adjusted | | | |
| --- | --- | --- | --- | --- | --- | --- | --- | --- | --- |
|  | **TKR** | | | | | | | | |
|  | HR | 95% CI | | *p*-value |  | HR | 95% CI | | *p*-value |
| *Medial compartment* |  |  |  |  |  |  |  |  |  |
| 1st quartile | 2.58 | 1.92 | 3.46 | **3.14 x10^-10^** |  | 2.44 | 1.77 | 3.38 | **6.58 x10^-08^** |
| 2nd quartile | 1.21 | 0.87 | 1.70 | 0.26 |  | 1.23 | 0.87 | 1.76 | 0.25 |
| 3rd quartile | 1.03 | 0.73 | 1.47 | 0.85 |  | 1.04 | 0.73 | 1.48 | 0.83 |
| 4th quartile | 1 |  |  |  |  |  |  |  |  |
| *Lateral compartment* |  |  |  |  |  |  |  |  |  |
| 1st quartile | 0.69 | 0.52 | 0.91 | **9.09 x10^-03^** |  | 0.62 | 0.45 | 0.85 | **3.20 x10^-03^** |
| 2nd quartile | 0.51 | 0.38 | 0.70 | **2.20 x10^-05^** |  | 0.48 | 0.34 | 0.66 | **9.45 x10^-06^** |
| 3rd quartile | 0.81 | 0.62 | 1.05 | 0.11 |  | 0.77 | 0.59 | 1.02 | 0.07 |
| 4th quartile | 1 |  |  |  |  | 1 |  |  |  |
| *Binary Q1 vs >Q1* | 2.38 | 1.93 | 2.93 | **3.51 x10^-16^** |  | 2.21 | 1.76 | 2.76 | **4.28 x10^-12^** |
|  | **HES-kOA** | | | | | | | | |
|  | OR | 95% CI | | *p*-value |  | OR | 95% CI | | *p*-value |
| *Medial compartment* |  |  |  |  |  |  |  |  |  |
| 1st quartile | 1.38 | 1.19 | 1.59 | **9.89 x10^-06^** |  | 1.54 | 1.31 | 1.81 | **1.13 x10^-07^** |
| 2nd quartile | 0.99 | 0.85 | 1.15 | 0.91 |  | 1.11 | 0.94 | 1.30 | 0.22 |
| 3rd quartile | 0.93 | 0.80 | 1.09 | 0.36 |  | 0.99 | 0.84 | 1.15 | 0.85 |
| 4th quartile | 1 |  |  |  |  | 1 |  |  |  |
| *Lateral compartment* |  |  |  |  |  |  |  |  |  |
| 1st quartile | 0.69 | 0.60 | 0.80 | **5.26 x10-^07^** |  | 0.79 | 0.67 | 0.93 | **4.63 x10^-03^** |
| 2nd quartile | 0.63 | 0.54 | 0.73 | **6.25 x10^-10^** |  | 0.69 | 0.59 | 0.81 | **4.15 x10^-06^** |
| 3rd quartile | 0.78 | 0.68 | 0.90 | **4.46 x10^-04^** |  | 0.81 | 0.71 | 0.94 | **4.67 x10^-03^** |
| 4th quartile | 1 |  |  |  |  | 1 |  |  |  |
| *Binary Q1 vs >Q1* | 1.41 | 1.26 | 1.58 | **1.34 x10^-09^** |  | 1.48 | 1.31 | 1.67 | **1.73 x10^-10^** |

When examining the association between quartiles of minimum joint space width (mJSW) and the risk of total knee replacement (TKR) and hospital diagnosed knee osteoarthritis (HES-kOA), we designated quartile 4 as the reference category (i.e., the greatest mJSW). Odds ratios (ORs) and hazards ratios (HRs) represent the difference in risk for someone in the first, second or third quartile versus the fourth quartile. For the binary mJSW variable, the HRs and ORs quantify the risk variation associated with values above the first quartile in the medial compartment, in comparison with the first quartile (representing the “unhealthiest” mJSW). CI, 95% confidence interval; Q1, first quartile of mJSW (medial compartment). p<0.05 are shown in bold.

Supplementary Table 9: Prevalence of manually graded osteophytes by site (n=6,719).

|  | **Medial femur** | |  | **Lateral femur** | |  | **Medial tibia** | |  | **Lateral tibia** | |
| --- | --- | --- | --- | --- | --- | --- | --- | --- | --- | --- | --- |
| *Grade* | *N* | *%* |  | *N* | *%* |  | *N* | *%* |  | *N* | *%* |
| 0 | 4812 | 71.62 |  | 6218 | 92.54 |  | 5126 | 76.29 |  | 4399 | 65.47 |
| 1 | 1538 | 22.89 |  | 292 | 4.35 |  | 1414 | 21.04 |  | 2136 | 31.79 |
| 2 | 264 | 3.93 |  | 133 | 1.98 |  | 150 | 2.23 |  | 147 | 2.19 |
| 3 | 105 | 1.56 |  | 76 | 1.13 |  | 29 | 0.43 |  | 37 | 0.55 |

Supplementary Table 10: Results of the unadjusted regression analysis examining the association of osteophyte grades with TKR and HES-kOA (n=6,719).

|  | **TKR** | | | |  | **HES-kOA** | | | |
| --- | --- | --- | --- | --- | --- | --- | --- | --- | --- |
|  | HR | 95% CI | | *p*-value |  | OR | 95% CI | | *p*-value |
| **Medial femur** | |  |  |  |  |  |  |  |  |
| grade 1 | 2.26 | 1.38 | 3.70 | 1.22 x10^-03^ |  | 1.51 | 1.16 | 1.98 | 2.50 x10^-03^ |
| grade 2 | 12.76 | 7.70 | 21.14 | 4.84 x10^-23^ |  | 7.30 | 5.25 | 10.15 | 3.36 x10^-32^ |
| grade 3 | 27.05 | 15.74 | 46.49 | 7.75 x10^-33^ |  | 10.60 | 6.76 | 16.61 | 7.52 x10^-25^ |
| **Lateral femur** | |  |  |  |  |  |  |  |  |
| grade 1 | 3.06 | 1.58 | 5.94 | 9.35 x10^-04^ |  | 2.29 | 1.51 | 3.46 | 9.07 x10^-05^ |
| grade 2 | 12.46 | 7.34 | 21.17 | 1.07 x10^-20^ |  | 8.34 | 5.58 | 12.46 | 4.43 x10^-25^ |
| grade 3 | 16.81 | 9.30 | 30.39 | 9.45 x10^-21^ |  | 6.47 | 3.72 | 11.26 | 3.72 x10^-11^ |
| **Medial tibia** | |  |  |  |  |  |  |  |  |
| grade 1 | 4.36 | 2.84 | 6.67 | 1.38 x10^-11^ |  | 3.23 | 2.56 | 4.08 | 8.89 x10^-23^ |
| grade 2 | 15.98 | 9.04 | 28.25 | 1.51 x10^-21^ |  | 7.16 | 4.64 | 11.05 | 5.79 x10^-19^ |
| grade 3 | 45.12 | 21.08 | 96.56 | 9.97 x10^-23^ |  | 21.09 | 9.91 | 44.87 | 2.47 x10^-15^ |
| **Lateral tibia** | |  |  |  |  |  |  |  |  |
| grade 1 | 3.42 | 2.22 | 5.28 | 2.55 x10^-08^ |  | 2.30 | 1.82 | 2.89 | 2.20 x10^-12^ |
| grade 2 | 11.60 | 5.99 | 22.45 | 3.62 x10^-13^ |  | 7.16 | 4.62 | 11.10 | 1.38 x10^-18^ |
| grade 3 | 49.30 | 24.91 | 97.57 | 4.55 x10^-29^ |  | 17.73 | 8.94 | 35.16 | 1.84 x10^-16^ |

The reference category for the regressions was set as grade 0. Abbreviations: CI, 95% confidence interval; HR, hazard ratio; OR, odds ratio.

Supplementary Table 11: Associations of osteophyte scores with TKR and HES-kOA (n=6,719).

|  | Unadjusted | | | |  | Adjusted | | | |
| --- | --- | --- | --- | --- | --- | --- | --- | --- | --- |
|  | **TKR** | | | | | | | | |
|  | HR | 95% CI | | p-value |  | HR | 95% CI | | *P-*value |
| *OP score* |  |  |  |  |  |  |  |  |  |
| 1 | 1.66 | 0.78 | 3.53 | 0.19 |  | 1.48 | 0.69 | 3.15 | 0.31 |
| 2 | 5.05 | 2.69 | 9.49 | **4.93 x10^-07^** |  | 3.98 | 2.11 | 7.52 | **1.98 x10^-05^** |
| 3 | 28.25 | 15.65 | 50.97 | **1.34 x10^-28^** |  | 18.30 | 9.98 | 33.57 | **5.77 x10^-21^** |
|  | **HES-kOA** | | | | | | | | |
|  | OR | 95% CI | | p-value |  | OR | 95% CI | | *P*-value |
| *OP score* |  |  |  |  |  |  |  |  |  |
| 1 | 1.74 | 1.25 | 2.41 | **1.01 x10^-03^** |  | 1.62 | 1.16 | 2.25 | **0.004** |
| 2 | 3.11 | 2.29 | 4.23 | **4.49 x10^-13^** |  | 2.72 | 1.99 | 3.71 | **<0.001** |
| 3 | 11.56 | 8.39 | 15.91 | **<0.0001** |  | 9.10 | 6.54 | 12.67 | **<0.001** |

The osteophyte scores were determined based on the cumulative sum of the four individual osteophyte grades: 0 (sum = 0), 1 (sum = 1), 2 (sum = 2-3), and 3 (sum = 4 or greater). In analysing the association between osteophyte scores and the probability of total knee replacement (TK)R and hospital diagnosed knee osteoarthritis (HES-kOA), the reference category was established as 0. The hazard ratios (HRs) and odds ratios (ORs) quantify the change in risk for individuals with scores of 1, 2, or 3 relative to those with a score of 0. Abbreviations: CI, 95% confidence interval.

Supplementary Figure 1: An example DXA image with the 129-point SSM template applied.

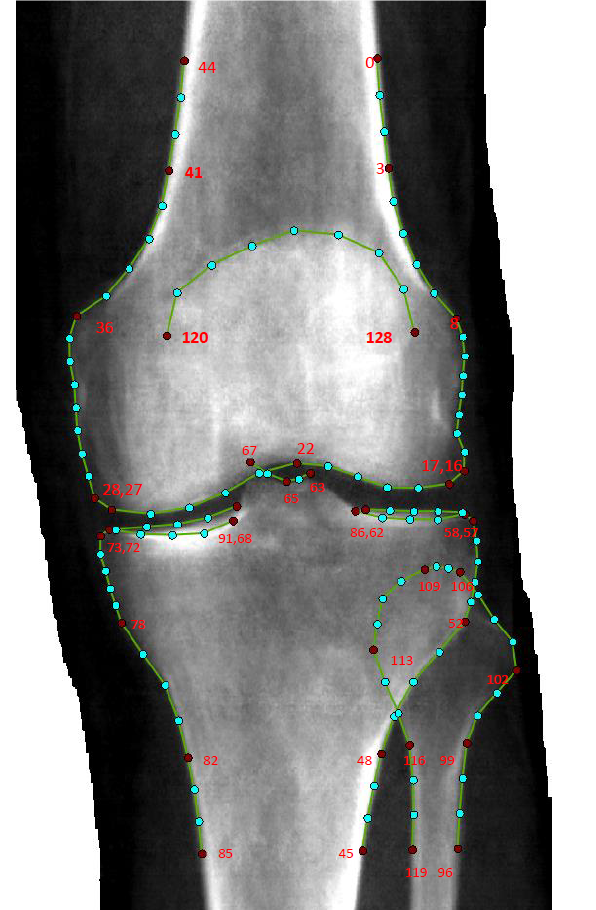

The image captured by the iDXA scanner displays the outline of a left knee and is overlapped by the statistical shape model (SSM) template. Landmark points, indicated in red, are placed on distinctive anatomical features and the remaining points are evenly distributed between each pair of landmarks in order to describe the shape.

Supplementary Figure 2: A graphical representation of the variance explained by each statistical shape model mode.

% variance explained

| Mode | Variance | Cumulative Variance |
| --- | --- | --- |
| 1 | 24.9539 | 24.9539 |
| 2 | 17.2009 | 42.1549 |
| 3 | 11.3261 | 53.481 |
| 4 | 7.25092 | 60.7319 |
| 5 | 4.80048 | 65.5324 |
| 6 | 4.01732 | 69.5497 |
| 7 | 2.77078 | 72.3205 |
| 8 | 2.68864 | 75.0091 |
| 9 | 2.41671 | 77.4258 |
| 10 | 2.1158 | 79.5416 |
| 11 | 2.01498 | 81.5566 |
| 12 | 1.57154 | 83.1281 |
| 13 | 1.55114 | 84.6793 |
| 14 | 1.25377 | 85.933 |
| 15 | 1.19653 | 87.1296 |
| 16 | 1.15625 | 88.2858 |
| 17 | 0.957874 | 89.2437 |
| 18 | 0.91166 | 90.1554 |
| 19 | 0.807257 | 90.9626 |
| 20 | 0.668533 | 91.6311 |
| 21 | 0.626208 | 92.2573 |
| 22 | 0.605768 | 92.8631 |
| 23 | 0.591817 | 93.4549 |
| 24 | 0.523583 | 93.9785 |
| 25 | 0.464259 | 94.4428 |
| 26 | 0.424681 | 94.8675 |
| 27 | 0.370638 | 95.2381 |

The proportion of variance explained by each mode is displayed on the y-axis and the number of modes of variation on the x-axis. The blue line shows the decreasing variance explained by individual modes, while the orange line displays the cumulative variance explained as additional modes are included. We used the first ten modes as these each contain more than 2% of the variance.

Supplementary Figure 3: Distribution of B-scores of healthy and diseased examples (n=37,843).

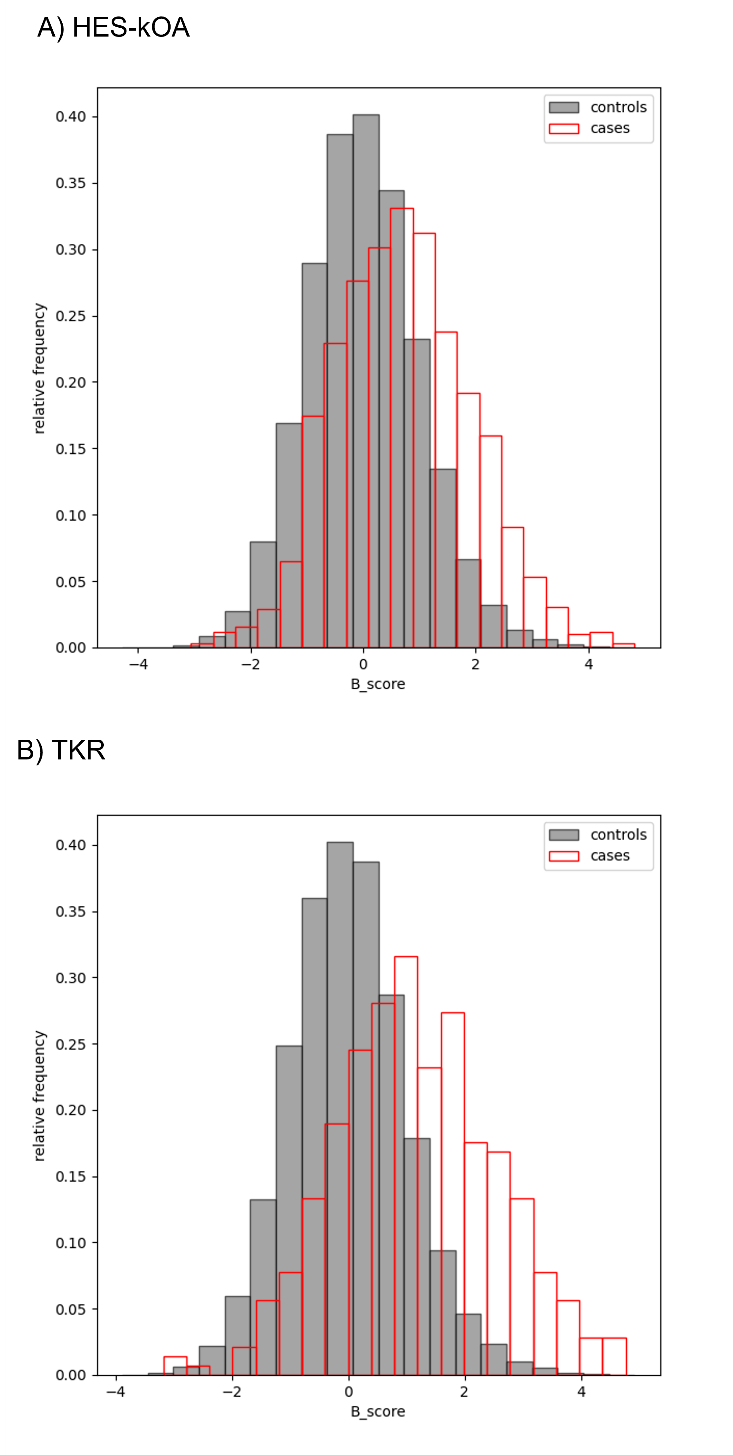

The plots depict the relative frequency of B-scores for healthy individuals in comparison to: A) hospital knee osteoarthritis (HES-kOA), and B) total knee replacement (TKR).

**Supplementary Methods**

*Statistical shape modelling*

SSM is a technique used to analyse and quantify shape variations in objects, such as anatomical joints. The technique involves applying a set of landmark points, which correspond to specific characteristics of the object, such as joint surfaces, to a sample of training images. The annotated points are used to align the shapes across all images. This is done using a Procrustes analysis, which adjusts the shapes to a common coordinate system by scaling, rotating, and translating them, thereby eliminating the impact of overall size. Once aligned, Principal Component Analysis (PCA) is employed to identify and quantify independent modes of shape variation (3, 4).

In this study, an initial training sample of 2000 left knee DXA scans, which was further extended by an additional 5000 scans, was selected to train an automated search model for point placement.  Among these, 20% were randomly chosen from individuals who self-reported non-specific osteoarthritis (OA). This enrichment facilitated effective learning of OA patterns by the model, a method previously implemented in similar studies (5, 6). The remaining 80% were randomly selected, while ensuring an equal sex distribution. Following this selection process, a total of 280 images were excluded from the analysis, leaving 6,720 DXA images. Of these exclusions, 105 images originated from the initial sample of 2000, while the remaining 175 images were part of the extension sample of 5000 images. The criteria for exclusion included poor image quality, significant metalwork, short femoral or tibial shafts, or participant withdrawal of consent. Additionally, two further images were subsequently excluded from the training set but they were retained within the broader analytic dataset. The first image featured a minor amount of metalwork, while the second displayed an unusually large tibial shaft. Consequently, the final training set comprised a total of 6,718 images.

A template comprising 129 landmark points (including 46 key points) was developed to outline the distal femur, proximal tibia, proximal fibula, and superior patella, excluding any osteophytes (Supplementary Figure 1). Our template's construction was informed by previously published knee shape models (7-10). Initially, the template included the medial femoral condyle, as in the Haverkamp model (9), which was developed using weight-bearing anteroposterior (AP) radiographs, however, we found that it was not consistently observable on the DXA images, which were taken in the supine position. It was therefore removed from the model during the early stages of development. Similarly, upon analysing a subset of images, we observed that only the top part of the patella was consistently visible. As a result, our template only incorporated landmark points on the superior region of this structure. Trained annotators marked up images from the training sample with these designated points.

We employed a 3-fold cross-validation model to assess the performance of the final search model, which was trained on the dataset of 6,718 images. This evaluation revealed that the mean point-to-point error was under 3mm for 95% of the images. Subsequently, we applied automated point placement to the remaining DXA images (n=31,207 after exclusions) using BoneFinder® (11), which employs a random-forest based algorithm. After point placement, a quality-of-fit score (QoF) was calculated for each image to measure how well the model matched to the image. These QoF scores were employed as a tool to visually inspect the images. Where necessary, trained annotators (RB and FS) manually refined the point placements to enhance the precision of the SSM. The final SSM model was built on 37,927 DXA images, which included 6,720 images from the initial training set and the remaining 31,207 available images. This produced a set of orthogonal modes of variation known as principal components (knee shape modes [KSMs]), which together explain 100% of variance in the data set. The first KSM corresponds to the highest proportion of variance, while subsequent KSMs represent diminishing amounts of variance (Supplementary Figure 2).

*Generation of Overall B-Score and Sex-Specific B-Scores*

To quantify the shape variations in knee joints, we employed the methodology introduced by Bowes et al. (6) to generate an overall knee shape variable termed the B-score. Using the shape parameters extracted from the SSM, we computed the mean shape for two distinct populations: a “kOA group” and a “healthy group”. The kOA group encompassed knees with either total knee replacement (TKR) or hospital-diagnosed knee osteoarthritis (HES-kOA), contingent upon the specific outcome being examined. The healthy group consisted of knees that did not progress to subsequent TRK or HES-kOA. We constructed a ‘kOA vector’, defined as the line passing through the mean shape of the kOA group and the healthy group. Each parameterized knee bone shape was then projected orthogonally onto this OA vector. The standard deviation (SD) of the projections from healthy cases was calculated and used to normalize the projections, resulting in the generation of the B-scores.

In the primary analysis, B-scores were determined for the entire population, i.e., males and females combined. As a sensitivity analysis, sex-specific B-scores were also calculated to explore potential differences in knee shape deviations between males and females. Separate sets of knee shape data were considered for each sex, and the calculations followed the same steps as described above for the overall B-score. Specifically, for each sex, we calculated the direction vectors based on the difference between the mean cases and the control shapes specific to that sex and divided the projections of knee shapes by the sex-specific standard deviations obtained from the healthy group.

*Automated generation of minimum joint space width (mJSW)*

A custom Python script was developed to automate the measurement of the minimum joint space width (mJSW) of the medial and lateral compartments. This script employs the shape modelling points located on the distal femur (medial points 24-30; lateral points 15-20) and proximal tibia (medial points 68-73; lateral points 57-62). We divided the mJSW in each compartment into quartiles and generated a binary variable that denoted whether the mJSW fell within the first quartile on the medial side. The first quartile corresponds to the smallest mJSW value.

*Creating a DXA-based osteophyte grade*

Manual grading of osteophytes was conducted in a subset of 6,719 images from the initial search model’s training set. As described above, this initial subset was derived from a pool of 7,000 DXA images; however, 280 were excluded from the analysis. Subsequently, one participant withdrew from the study, necessitating their removal from the analytic dataset. Each image was visually assessed for osteophytes on a 0-3 scale, referencing a DXA-based atlas created by RB and FS (supplementary doc) with input from DW (see acknowledgements). The evaluation specifically focused on four sites on the lateral and medial aspects of the femur and tibia. Intra-observer repeatability was assessed on a random sample of 200 images, demonstrating good agreement (κ = 0.80). Osteophyte severity across the whole knee joint was captured by summing osteophyte grades in all four locations, resulting in a value ranging from 0 - 12 (Supplementary Table 4). A score was then assigned based on the sum as follows: 0 (sum = 0), 1 (sum=1), 2 (sum = 2-3), 3 (sum =4 or greater). Similar methods have been used in previous studies on the hip and knee (12-14). This categorisation reflects the observation that the number of knees with a sum of 4 or more across all sites was relatively low.
